# Cognitive Intervention Effects Vary as a Function of Plasma Neurofilament Light Chain Levels: A PICMOR Randomized Controlled Trial

**DOI:** 10.1101/2023.03.30.23287976

**Authors:** Mihoko Otake-Matsuura, Hikaru Sugimoto, Takuya Sekiguchi, Masato S. Abe, Kumi W. Miura, Seiki Tokunaga, Shoshin Akamine, Taishiro Kishimoto, Takashi Kudo

## Abstract

**Background:** There is a difference in the neuronal state of individuals. However, this has not been taken into consideration in most intervention studies. Recent advances in analytical technologies in hematological examination enabled us to evaluate neuronal states in a relatively convenient manner.

**Objective:** Using these advanced technologies, we aimed to investigate whether cognitive intervention effects vary as a function of levels of blood-based biomarkers, such as neurofilament light chain (NfL), since plasma NfL could be a biomarker of neurodegenerative diseases including Alzheimer’s disease.

**Methods:** In this study, we employed a group conversation-based intervention methodology named Photo-Integrated Conversation Moderated by Robots (PICMOR), which has been shown to improve verbal fluency in older adults. To examine the possibility of varying effects of this intervention method according to the neuronal state of each individual, we conducted a randomized controlled trial (UMIN Clinical Trials Registry number: UMIN000036599) and investigated how longitudinal changes in cognitive performance, such as verbal fluency, vary with the NfL level measured at the baseline.

**Results:** As the main result, positive intervention effects of PICMOR on verbal fluency were observed in individuals with lower level of NfL, which indicate a relatively intact neuronal state, whereas negative intervention effects were identified in individuals with higher NfL level.

**Conclusion:** Our findings suggest that cognitive intervention effects vary depending on level of Nfl in the plasma. Thus, future intervention studies should take into account the neuronal status of the participants to examine intervention effects.

## INTRODUCTION

Dementia is one of the major issues in an aging global population, and its prevention would yield great social benefits. The number of people living with dementia has rapidly increased to 50 million, particularly because of aging populations in low and middle-income countries [1]. Alzheimer’s disease (AD) is the most common form of dementia, and its pathology progresses for a long time before symptoms appear. AD continuously encompasses several phases: a phase in which pathology is present despite cognitive normality [2], a mild cognitive impairment (MCI) phase [3], and, a dementia phase [4]. Since the development of therapeutic agents has not been successful because of the progressive nature of AD, prevention of AD attracts much more attention to delay the progress of pathology or cognitive decline at the normal, preclinical, or MCI phase. Based on the mechanism, cognitively normal older adults are diverse in terms of neuronal state related to AD. Some may have neuronal degeneration, some may be at higher risk of neuronal degeneration, and others may not. Therefore, it is possible that there is an individual difference in the degree of the effect of interventions on cognitive functions according to the neuronal state of each person.

Previously, the assessment of neuronal state was difficult since the measurement of pathology required positron emission tomography (PET) scanning or collection of cerebrospinal fluid (CSF). Recently, however, cutting-edge analytical technologies in hematological examination have enabled us to estimate neuronal state relatively easily. In this method, the plasma is separated from the blood, and the concentration of neurofilament light chain (NfL), a subunit of neurofilaments, in the plasma is measured, which indicates axonal degeneration. NfL is a neuronal cytoplasmic protein <u>that is</u> highly expressed in large-caliber myelinated axons [5]. In neurodegenerative diseases, such as AD, the disruption of the axonal membrane causes a sharply increased release of NfL into the interstitial fluid, followed by its release into CSF and blood [6]. Even in normal populations, NfL is constantly released from axons, and the release level is increased with advancing age [7, 8]. Thus, the concentration of this protein in CSF and blood can be considered as an indicator of axonal injury. In recent studies, blood-based NfL measurement has been widely adopted because of the following reasons: it is less invasive and more convenient relative to the CSF-based method; NfL levels measured by the two methods are correlated [9-12]; and a more sensitive assay, called single-molecule array (Simoa), has been developed [13]. In the present study, we used blood-based method to measure NfL levels and regarded NfL levels as a reliable index of neuronal state, as it has been hypothesized to be a nonspecific marker of neurodegeneration [9-12, 14]. In addition, we employed other blood-based biomarkers as references for comparisons: amyloid-β (Aβ) (Aβ40 and Aβ42) in extracellular vesicles (EV).

The aim of this study was to examine whether cognitive intervention effects in healthy older adults vary according to their neuronal state. We employed conversational interventions to accumulate evidence on the effects of interventions based on social activities, which is still limited in available literature [15]. One of the reasons that social activity-based intervention is scarce is that social activities, such as conversations, occur spontaneously, and it is difficult to regulate their qualities and quantities. To address this issue, we previously developed a cognitive intervention program that utilizes group conversations named Photo Integrated Conversation Moderated by Robots (PICMOR) [16]. By using a robot system, controlling the quantity of conversation is enabled. The system contains two key features: (i) a robot moderator that controls the turn-taking so that all participants speak and listen for the same length of time, and (ii) photos taken by the participants are shown and discussed sequentially with the announcement of a robotic moderator. These features are important for exercising executive functions and episodic memory because the participants have to speak within a designated period of time and have to ask and answer questions with flexibility; thus, they would be trained to organize their thoughts and produce them as words. The participants are required to recall their autobiographical memories using the photos, and therefore, their ability to recall from memory would be also exercised. Indeed, we previously identified the beneficial effects of the PICMOR program on verbal fluency in older adults [16].

In the present study, we investigated whether the effect of the conversation-based intervention on cognitive functions differs depending on the neuronal state of each participant estimated from blood biomarkers. To do this, we conducted a randomized controlled trial (RCT) on healthy older adults, assuming that the degree to which sub-domains of cognitive functions involved in conversations were enhanced by the PICMOR program would vary from individual to individual as a function of the blood levels of NfL. More specifically, individuals with lower levels of NfL, which imply a relatively intact neuronal state, would show larger improvement in cognitive performance, such as verbal fluency [16, 17]. This is because the intact neural basis can play its role and enjoy the benefits of cognitive training.

## METHODS

### Study design and procedure

The study was a randomized control trial conducted between May and September 2019 (UMIN Clinical Trials Registry number: UMIN000036599). Figure 1 describes the CONSORT diagram showing an overview of the current study. First, 70 participants were assessed for eligibility by medical interview, cognitive tests, and radiologic interpretations. Eighteen participants were excluded under our exclusion criteria and one participant withdrew their consent. Therefore, 51 participants were included in the randomization. After baseline assessment, block stratified randomization was performed according to scores of the Japanese version of the Mini-Mental State Examination (MMSE-J) [18] and the Montreal Cognitive Assessment (MoCA-J) [19], and randomly assigned participants to either the intervention (*n* = 24) or control group (*n* = 27). We divided participants in the intervention group into six subgroups of four. The intervention group was given 30-minute weekly intervention session, followed by 30-minute explanation of the intervention. The control group visited the office weekly as an active control session. During the intervention period, two participants were lost to follow-up, with one individual belonging to the intervention group and the other from the control group. Subsequent to the 12-week intervention period, post-assessment was carried out for 49 subjects (intervention: *n* = 23, control: *n* = 26). This study was approved by the Institutional Review Board. Written informed consent was obtained from all participants.

**Fig. 1.**
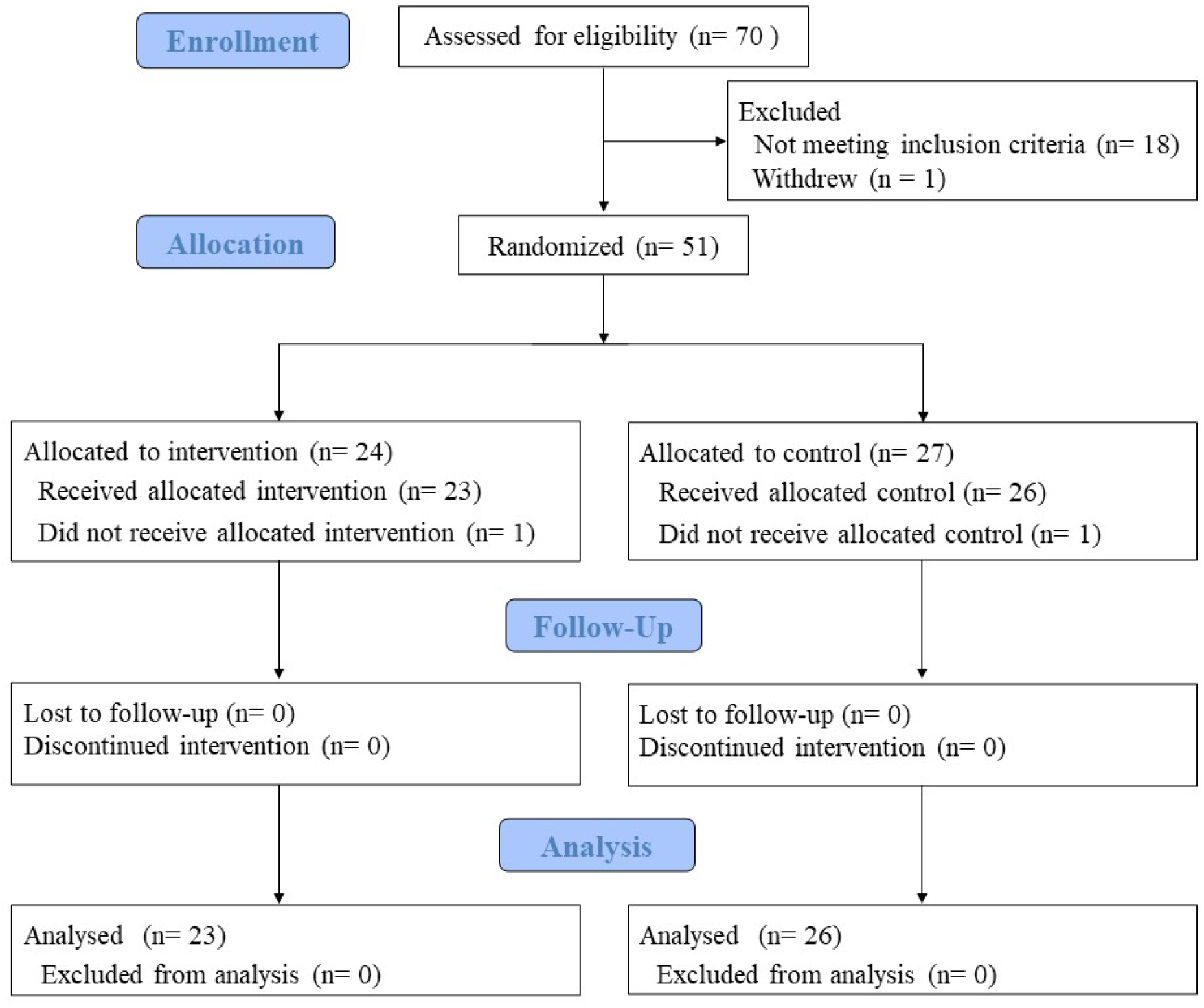
The CONSORT diagram of the current study.

### Participants

The participants were older adults over 65 years in community living who were enrolled from Silver Human Resources Center, Japan. The exclusion criteria of the current study were the following: participants with a score of MMSE-J less than 24, neurological impairment, dementia, and any medical condition or medication with a known effect on the central nervous system. Radiological findings assessed by a medical doctor using multiple magnetic resonance imaging (MRI) data, including T1-weighted, T2-weighted, and FLAIR images, were also used as exclusion criteria: cerebral infarctions, meningeal tumors, hemorrhage, or abnormal signals probably caused by metal artifacts.

### Neuropsychological examination

Outcome measurements of cognitive performance in the current study were conducted using MoCA-J, MMSE-J, semantic verbal fluency test [20], subtests of the Wechsler Memory Scale-Revised (WMS-R) [21], including logical memory I and II, subtests of the Wechsler Adult Intelligence Scale-Fourth Edition (WAIS-IV), including the digit symbol coding and digit span forward and backward tests [22], and Trail Making Test (TMT) [23].

MoCA-J and MMSE-J are brief cognitive screening tools composed of multiple domains of cognitive function. In this study, the total score in each test was calculated.

Verbal fluency was evaluated by two distinct tests: the phonological verbal (or letter) fluency test from MoCA-J and semantic verbal (or category) fluency test. In the letter fluency test, we instructed to participants to generate as many words as possible in 1 min, with each word being started by the Japanese character ‘ka’. In the category fluency test, participants were required to speak as many words as they could belonging to the category of animals in 1 min. The two types of verbal fluency tasks differ in whether words are generated in a phonologically or semantically driven way. The overall count of words produced was recorded in each test.

Logical memory I and II are measure of narrative episodic memory that assess both immediate and delayed verbal memory. After reading a brief story, the examiner instructed the participants to reproduce the story’s content immediately and again after a duration of 30 minutes. Scores were calculated by the sum of the number of elements that were recalled. To avoid learning effects, we used different test stories at the baseline assessment and post-assessment in this study.

The processing speed and working memory are assessed by the digit symbol coding test. In this test, the participants were instructed to transcribe as many of the corresponding symbol as possible, followed by a list of digits. Simple memory span and working memory capacity are measured by the digit span forward and backward tests, respectively. The examiner told the sequence of numbers and required participants to recall the sequence forward and backward.

TMT assesses attention and executive function of individuals. In this study, TMT was conducted using a PC. In TMT-A, a series of circles, numbered consecutively from 1 to 25, were randomly dispersed throughout the visual display, and the participants were asked to click the cursor on each circle in numerical order. In TMT-B, a series of circles included not only numbers ranging from 1 to 13, but also the initial 12 Japanese Hiragana character. We instructed to click on numbers and letters alternatively. The time to perform these tasks was recorded.

### Blood sampling and separation of plasma

Blood was collected in EDTA tubes in the Advanced Imaging Center (AIC) Yaesu Clinic, Tokyo, Japan. The blood was centrifuged at 2,500 g for 15 min at room temperature, 18 °C, within 24 h after sampling, and stored at -80 °C for future analyses and transported to Osaka University. The following procedures, isolation of neuron-derived extracellular vesicles (EV) (NDE) fraction from plasma, measurements of Aβ40, Aβ42 in plasma-derived NDE fraction, and measurements of serum NfL were conducted at Osaka University, Osaka, Japan.

### Measurements of serum NfL

We measured Plasma NfL concentration twice with the Simoa platform, using Simoa NF-Light Advantage kits on a Simoa HD-1 Analyzer instrument (Quanterix Corporation) [24]. We set 0.466 pg/mL as the lower limit for NfL concentration. We excluded samples with either fatal measurement errors or coefficients of variance (CV) > 20 %. In all batches, we measured two types of quality-control samples provided in the kit twice, in order to ensure measurement validity. The mean CV of duplicates was 4.4 %.

### Isolation of NDE fraction from plasma

NDE fractions were isolated from plasma using the previous method reported by Goetzl and Kapogiannis group [25-27] with some modifications. 7.5 μL thrombin (Thrombin Plasma prep #TMEXO-1; System Bioscience, CA, USA) was added to 250 μL plasma and incubated for 30 min at room temperature to remove fibrin by clot formation. After the addition of 242.5 μL of Dulbecco’s phosphate-buffered saline with no calcium and magnesium ion (D-PBS(-)) (#14249-95; Nacalai Tesque, Kyoto, Japan), tubes were gently mixed by inverting three times, and centrifuged at 4,000 g for 20 min at 4 °C. Supernatants were transferred to fresh tubes and added 126 μL of ExoQuick solution (#EXOQ5A-1; System Bioscience, CA, USA), and gently mixed by inverting three times. Tubes were incubated for 60 min at 4 °C and centrifuged at 1,500 g for 20 min at 4 °C.

Supernatants were discarded after centrifugation, and the pellet was resuspended in 250 μL of deionized water. The solution was incubated overnight at 4 °C with rotation for complete resuspension. To isolate NDE fraction from total plasma-EVs, 2 μg of biotinylated anti-L1CAM antibody (#13-1719-82; eBioscience, CA, USA) in 21 μL of 3 % bovine serum albumin (BSA) (Blocker BSA, #37525; Thermo Fisher Scientific, MA, USA) diluted with D-PBS(-) was added and incubated for 60 min at 4 °C with rotation, followed by addition of 7.5 μL of streptavidin-agarose resin (Pierce Streptavidin Plus UltraLink Resin, #53116; Thermo Fisher Scientific, MA, USA) and 12.5 μL of 3 % BSA. The solution was further incubated for 30 min at 4 °C with rotation, centrifuged at 200 g for 10 min at 4 °C, then the supernatant was carefully removed. NDE fraction was eluted by 100 μL of 0.1 M glycine-HCl (pH 3.0), the resin was removed by centrifugation at 4,500 g for 10 min at 4 °C, followed by neutralization with 7.5 μL of 1 M Tris-HCl (pH 8.0) and 7.5 μL of 10 % BSA. EVs were lysed by adding 135 μL of M-PER (#78501; Thermo Fisher Scientific, MA, USA) and two freeze-thaw cycles.

All D-PBS(-), deionized water, and M-PER were supplemented with protease inhibitor cocktail (cOmplete Protease Inhibitor Cocktail, #04693116001; Sigma-Aldrich, MO, USA) and phosphatase inhibitor cocktail (PhosSTOP, #4906847001; Sigma-Aldrich, MO, USA) to three times the concentration recommended by the manufacturer, before use.

### Measurements of Aβ40, Aβ42 in plasma-derived NDE fraction

We measured Plasma-derived NDE Aβ40/Aβ42 concentration twice with the ultrasensitive Simoa platform, using Simoa Neurology 3-Plex A Advantage kits on a Simoa HD-1 Analyzer instrument (Quanterix Corporation) [24]. We diluted samples 4-fold with sample diluent reagent in the instrument during measurement. The lower limit of quantification for Aβ40 and was 0.675 pg/mL and that of Aβ42 was and 0.142 pg/mL. We excluded samples with either fatal measurement errors or CV > 20 % from the analysis. In all batches, we measured two types of quality-control samples provided in the kit twice, in order to ensure measurement validity. The mean CV of duplicates was 4.4 % and 3.5 %, respectively.

### MRI data acquisition and preprocessing

Anatomical brain structures were scanned using a high-resolution T1-weighted image. The scan parameters were as follows: repetition time = 6.41 ms, echo time = 3.00 ms, field of view = 24.0 cm × 24.0 cm, matrix size = 256 × 256, slice thickness/gap = 1.2/0 mm, and 170 sagittal slices. The participant’s head was fixed with a belt and foam pads during the scan. Resting-state functional images and diffusion tensor images were also collected and used for other purposes. All MRI data were acquired using a Philips Achieva 3T MRI scanner at the AIC Yaesu Clinic. The T1-weighted images were preprocessed using Statistical Parametric Mapping (SPM) 12. The images were segmented into intracranial parts of gray matter (GM), white matter (WM), CSF, and other non-brain structures using the standard unified segmentation module [28]. From the segmented images, individual GM, WM, and CSF volumes were calculated for analysis. Total brain volumes (TBVs) and total intracranial volumes (TIVs) were defined as the sum of GM and WM and the sum of the three parts (GM + WM + CSF), respectively.

### Covariates

Age, sex, educational level, family structure, competency, depressive tendency, and quality of life were assessed as covariates. Competency was assessed by the Tokyo Metropolitan Institute of Gerontology-Index of Competence (TMIG-IC) [29], and the subject’s depressive tendency was evaluated by the Geriatric Depression Scale short form (GDS-15) [30]. World Health Organization quality of life questionnaire 26 was used to evaluate the quality of life of the participants [31].

### Statistical analysis: Intervention effects

To estimate the intervention effect, we performed the linear mixed models with random intercepts for outcome measures (scores of cognitive tests, brain volumes, and levels of blood biomarkers) by using the R package ‘lme4’ [32]. The models included the group assignment factor (the intervention group was coded as 1 and the control group was coded as 0), the time factor (pre-experiment was coded as 1 and post-experiment was coded as 0), and their interaction term as independent variables. We reported the interaction term as the intervention effect.

### Statistical analysis: Intervention effects depending on levels of blood biomarkers

To observe at which biomarker level the time effect and the intervention effect on cognitive test scores conditional on the biomarker level were significant, we constructed the linear mixed models with random-intercepts, which included the group factor, the time factor, the biomarker levels, and all the possible interaction terms. We then applied the simple slope analysis to those models, in which we subtracted specific values from the raw values of biomarker levels and then conducted the same models again.

We reported ranges of biomarker levels where the time effects within the intervention and control groups and the intervention effect on the outcomes were significant at levels of 0.05.

## RESULTS

### Baseline characteristics of participants

Table 1 compares the baseline characteristics of study participants in the intervention and control groups.

**Table 1.** Baseline characteristic between intervention and control group.

|  | Intervention | Control |
| --- | --- | --- |
|  | (*n* = 23) | (*n* = 26) |
| Age (Mean ± SD) | 72.30 ± 4.15 | 72.04 ± 4.05 |
| Gender (Female; *n*, %) | 13 (56.5 %) | 17 (65.4 %) |
| Education (≥ 13 years; *n*, %) | 16 (69.6 %) | 21 (80.8 %) |
| MMSE (Mean ± SD) | 28.17 ± 1.75 | 28.19 ± 1.70 |
| GDS-15 (Mean ± SD) | 1.57 ± 1.65 | 2.65 ± 3.07 |
| TMIG-IC |  |  |
| Total score (Mean ± SD) | 12.2 ± 0.96 | 11.88 ± 1.53 |
| Instrumental activity of daily living (Mean ± SD) | 4.83 ± 0.49 | 4.88 ± 0.33 |
| Intellectual activity (Mean ± SD) | 3.74 ± 0.54 | 3.62 ± 0.64 |
| Social role (Mean ± SD) | 3.70 ± 0.47 | 3.38 ± 0.90 |

### Intervention effects in cognitive test scores, brain volumes, and blood biomarkers

Table S3 compares the cognitive test scores and brain volumes measured at pre- and post-experiment between the intervention and control groups. No statistically significant intervention effects were observed for any of the scores and brain volumes.

Table S4 compares the levels of blood biomarkers pre- and post-experiment between the intervention and control groups. There were no significant intervention effects observed on levels of any biomarkers.

### Intervention effects depending on levels of blood biomarkers

For lower (higher) levels of NfL, a significantly positive (negative) intervention effect on category fluency was observed (Fig. 2).

**Fig. 2.**
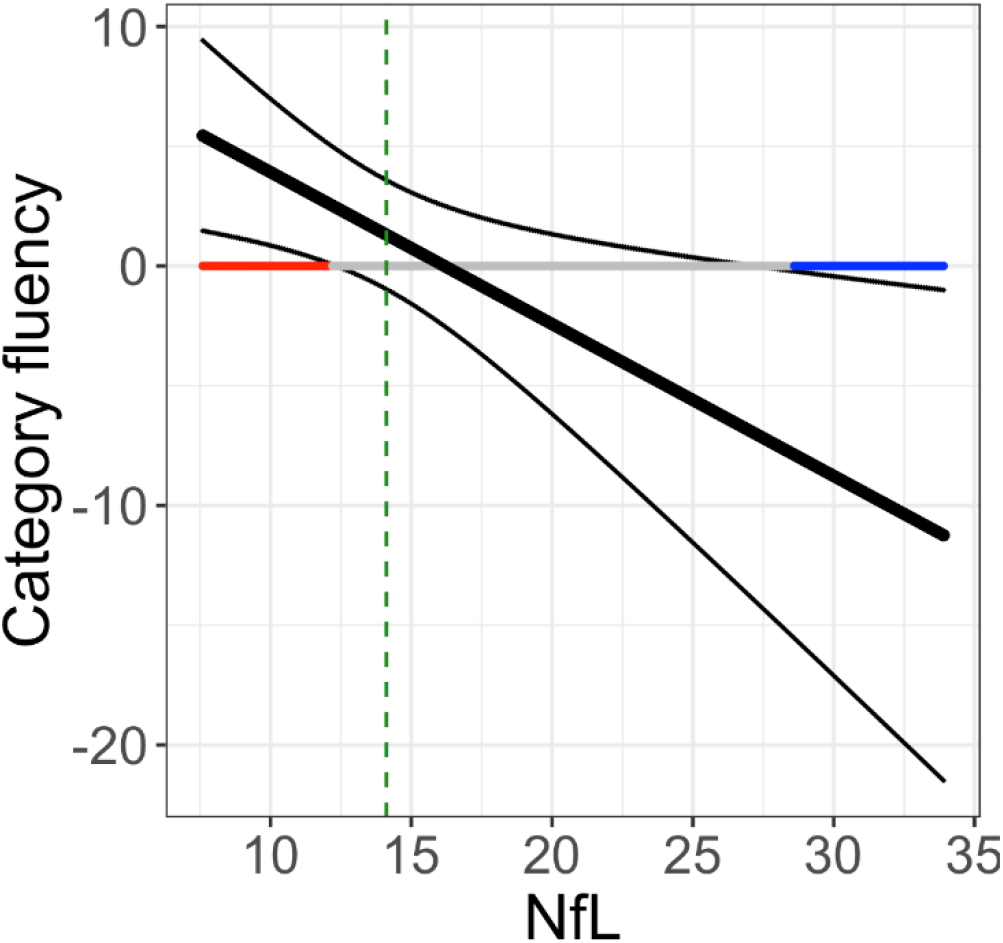
The intervention effects on the scores of category fluency tests for varying levels of NfL. The black thick line indicates the estimated intervention effect, and the black thin lines indicate the upper and lower bounds of the associated 95 % confidence interval. On the horizontal line at *y* = 0, the red region shows the region of the biomarker level in which the intervention effect was positively significant at the 5 % level, and the blue region shows the region of the biomarker level in which the intervention effect was negatively significant at the 5 % level. The gray region shows the region of non-significance for the intervention effect. The range of the horizontal axis is truncated to the observed ranges. The vertical dashed green line indicates the median of the observed level of the biomarker.

For a higher level of NfL, significantly negative intervention effects on logical memories I and II were observed (Fig. S1A and B), and for a level of NfL close to the median, a significantly negative intervention effect on Digit span backward was observed (Fig. S1C).

For a lower (higher) level of Aβ40, a significantly positive (negative) intervention effect on digit span forward was observed (Fig. S1D).

For a level of Aβ40 close to the median, a significantly negative intervention effect on digit span backward was observed (Fig. S1E). For a higher level of Aβ42, a significantly negative intervention effect on the volume of white matter was observed (Fig. S1F).

Figure S2 shows that the time effects on cognitive test scores within the intervention and control groups were not negative. By contrast, the time effects on brain volumes were negative in many cases.

## DISCUSSION

In this study, we examined whether cognitive intervention effects vary as a function of levels of blood-based biomarkers, assuming that individuals with lower levels of NfL would show a larger enhancement in verbal fluency. Although we did not find significant intervention effects in any task scores when the blood-based biomarkers were not taken into consideration, we successfully identified differential intervention effects on performance in one of two verbal fluency tasks when individuals were categorized by the level of NfL.

Our previous RCT employed a letter fluency task and identified a significant intervention effect of PICMOR on this task score [16], whereas the present RCT used a category fluency task as well as the letter one and found no significant intervention effects on both task scores (see Table S3). This difference could be explained by a difference in the baseline level of task performance between the previous and present RCT participants. In this study, both the intervention and control groups scored 14 on average in the letter fluency test, which are significantly higher than the previous participants who scored 11 on average [16]. Considering that normative data regarding this task performance show that people aged 60 and 70 years score 7 on average [33], the baseline level of participants in this study is extremely high, and hence it is possible that there was a smaller room for growth in the task performance compared to the previous RCT participants. Similarly, the current participants scored 17 on average at baseline in the category fluency test, which is extremely higher than normative data of people aged 60 and 70 years scoring the 14 and 11 on average, respectively [33]. Nevertheless, when considering NfL levels, we found a significantly positive intervention effect in the semantic verbal fluency task for individuals with lower levels of NfL (see Fig. 1A). In contrast, no significant effects were observed in the phonological one, even when the NfL was taken into consideration. This difference may be reasonable because the phonological one is more difficult to perform than the semantic one [33]. The present findings suggest that even in populations with high level performance, there is an individual difference in the neuronal state and cognitive intervention effects are prominent for individuals with a relatively intact neuronal status. On the other hand, a negative intervention effect was observed in those who had higher levels of NfL. Thus, the finding of no significant intervention effects on verbal fluency unless NfL levels are not taken into account could be also explained by the possibility that the effects were counterbalanced between individuals with lower and higher NfL levels. Taken together, our findings suggest that cognitive intervention effects vary as a function of NfL levels.

This study showed an advantage of using plasma NfL levels in intervention studies. Recently, there has been a growing interest in blood-based NfL as a biomarker of neurodegenerative diseases [9-12, 14]. In addition, several studies have attempted to use it as a measure of treatment for diseases [9]. In aging research, the NfL levels have been shown to increase with normal aging [7, 8]. This index has also been used in cohort and RCT studies for healthy older adults [34, 35]. For example, Desai et al. (2022) utilized the NfL levels to classify the participants and investigated the association between physical activity levels and cognitive decline [34]. Our study proposed a new usage of the NfL by employing it for the classification of participants to examine a cognitive intervention effect among healthy older populations. This is informative for other intervention studies because the effects of any intervention methods vary as a function of NfL levels. In contrast, we could not identify a similar pattern when using Aβ levels for classification. This is presumably because the measurement may not reflect only Aβ levels measured from neuron-derived exosomes. We originally used L1CAM as a marker of brain neuronal EV and collected exosomes with L1CAM to measure Aβ levels in neural cells. However, the utility of L1CAM as a marker of brain neuronal EVs has been questioned recently because some of L1CAM is expressed in other organs, such as the kidneys, and most of L1CAM in the blood is truncated, lacking the transmembrane region which anchors L1CAM to the membrane of EVs [36-38]. Thus, it is important to develop a new technology to distinguish neuron-derived exosomes from those that came out of other organs. Taken together, the plasma NfL measured in this study could be considered as purely reflecting neuronal state. In this study, significant negative intervention effects were observed on logical memory task scores (I and II) (Fig. S1) as well as the categorical verbal fluency task score among participants with higher levels of NfL (see Fig. 2). In addition, participants with middle levels of NfL showed significant negative intervention effects in the digit span backward test (see Fig. S1). These findings suggest that individuals with high or medium levels of NfL do not benefit from cognitive intervention due a relatively vulnerable neural basis. From the results, we can at least derive a practical implication that an appropriate intervention program should be designed according to the levels of biomarkers. Future research needs to investigate how strong the intervention strength should be to improve their cognitive performance by changing the frequency or duration of interventions or the cognitive load demanded in interventions. A cohort study reported that even in individuals with higher levels of NfL, those who have high or medium levels of physical activity showed reduced cognitive decline relative to those who have low physical activity levels [34]. Considering that the PICMOR method has various variables that can be changed, such as the number of topics to be provided, it may be effective to increase the cognitive demands during the intervention as well as to increase the frequency or duration of interventions for individuals with lower levels of NfL.

The PICMOR-based conversation is designed to train cognitive functions, such as executive functions and episodic memory [16]. In this structured group conversation, participants were required to make a presentation on recently encountered events within a limited time, ask and answer questions among group members, and refrain from making a comment when other members were talking. To provide topics, they had to recall their autobiographical memory and explain it in a concise manner. To ask questions, they had to temporarily retain information in working memory and understand what others meant and what was unclear with reference to one’s knowledge. As expected, our series of RCTs provided consistent evidence that the verbal fluency ability, which is based on executive control and verbal abilities [39], can be improved by engaging in the PICMOR-based conversation. However, it remains unclear what components in this structured conversation are responsible for the enhancement of verbal fluency [40]. Further research is needed to address this issue by investigating whether repeated training specialized in each component of conversations, such as asking questions, results in the improvement of cognitive abilities. Furthermore, considering the pandemic of COVID-19, it is important to develop more easily available methods for people in isolation or those who cannot go out, which do not require an on-site conversation, such as tablet-based or application-based conversations.

In conclusion, we found that cognitive intervention effects vary as a function of levels of blood-based biomarkers. Importantly, we demonstrated that the beneficial effects of our recently developed intervention program on verbal fluency are prominent in individuals with lower levels of NfL. From these findings, we can suggest for future intervention studies that it is important to estimate the neuronal state of participants when determining the effectiveness of intervention methodologies.

## Data Availability

All data produced in the present study are available upon reasonable request to the authors

## ACKNOWLEDGMENTS

We thank the staff of the AIC Yaesu Clinic for their technical assistance in the MRI scanning and blood sampling. This work was supported by JSPS KAKENHI (Grant Numbers: JP18KT0035, JP19H01138, JP19K14489, JP20H05022, JP20H05574, JP20K19471, JP22H04872, JP22H00544) and the Japan Science and Technology Agency (Grant Numbers: JPMJCR20G1, JPMJPF2101, and JPMJMS2237).

## SUPPLEMENTARY MATERIAL

**Fig. S1.**
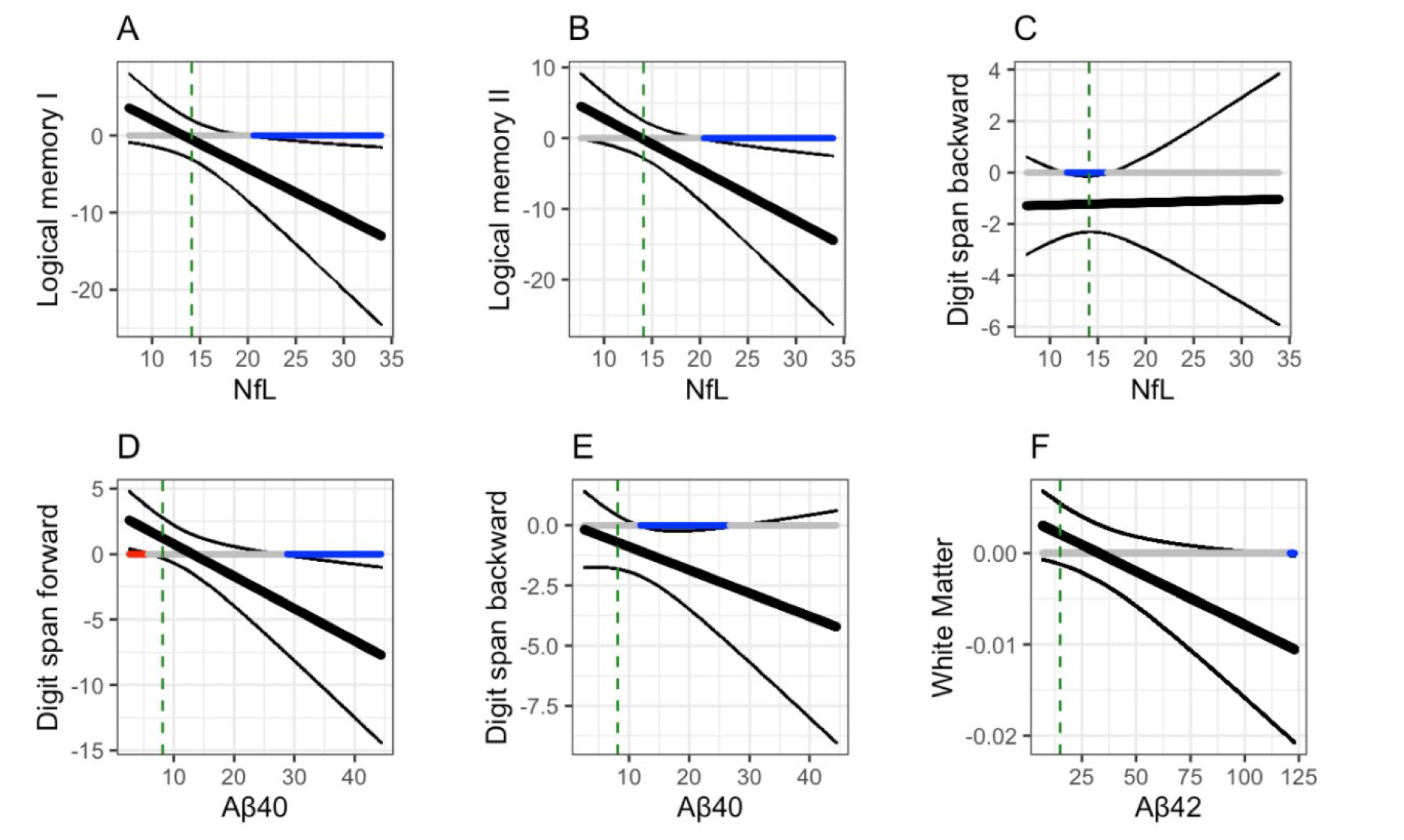
The intervention effects on the scores of the cognitive tests and brain volumes for varying levels of biomarkers. The black thick line indicates the estimated intervention effect, and the black thin lines indicate the upper and lower bounds of the associated 95 % confidence interval. On the horizontal line at *y* = 0, the red region shows the region of the biomarker level in which the intervention effect was positively significant at the 5 % level, and the blue region shows the region of the biomarker level in which the intervention effect was negatively significant at the 5 % level. The gray region shows the region of non-significance for the intervention effect. The range of the horizontal axis is truncated to the observed ranges. The vertical dashed green line indicates the median of the observed level of the biomarker.

**Fig. S2.**
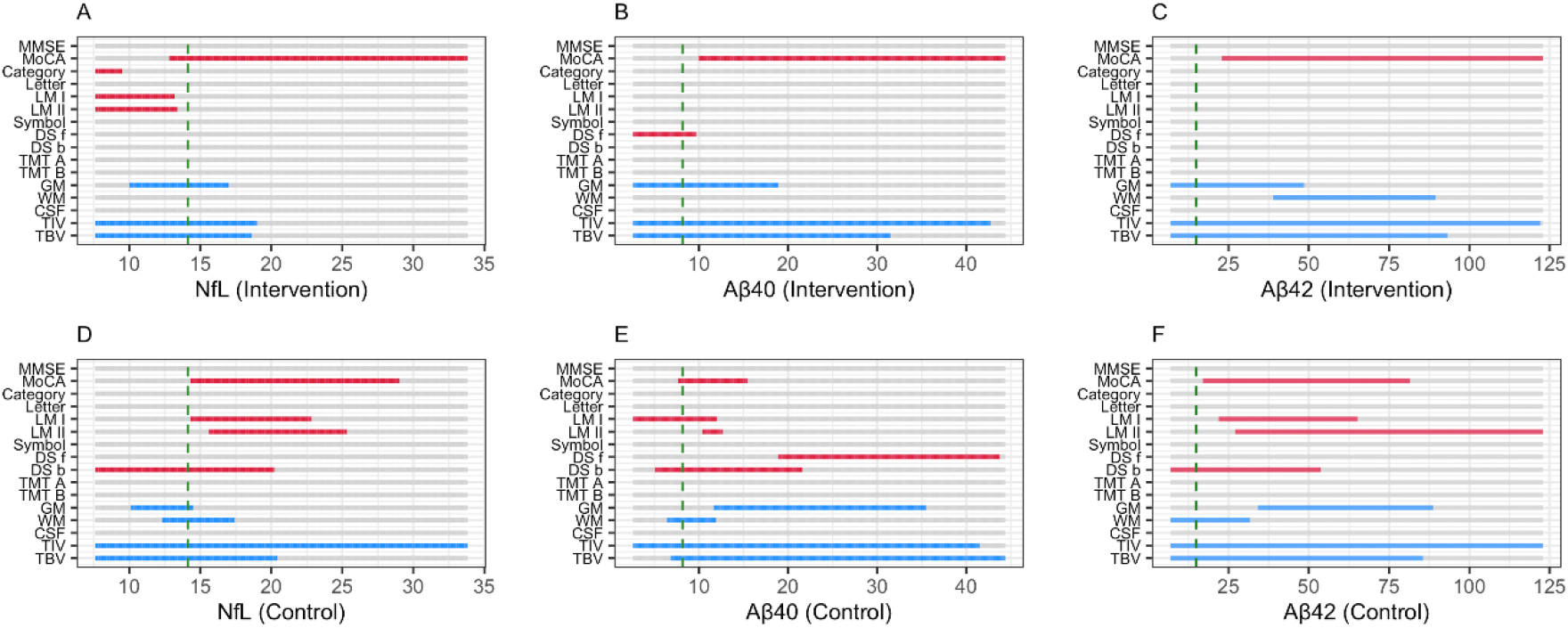
The time effects on the scores of the cognitive tests and brain volumes for varying levels of biomarkers. (**A-C**) show the results within the intervention group, and (**D-F**) show those within the control group. On each horizontal bar, the red (blue) region shows the region of the biomarker level in which the time effect was positively (negatively) significant at the 5 % level. Gray region shows the region of non-significance for the time effect. The range of the horizontal axis is truncated to the observed ranges. The vertical dashed green line indicates the median of the observed level of the biomarker.

**Table S3.** Comparison of cognitive test scores and brain volumes at pre- and post-experiments between the intervention and control groups.

|  | Intervention |  | Control |  | Estimates |  |  |
| --- | --- | --- | --- | --- | --- | --- | --- |
| | Pre Mean (SD) | Post Mean (SD) | Pre Mean (SD) | Post Mean (SD) | Time (SE, *p*) | Group (SE, *p*) | Time × Group (SE, *p*) |
| Cognitive test scores |  |  |  |  |  |  |  |
| MMSE-J | 28.174 (1.749) | 28.261 (1.322) | 28.192 (1.698) | 28.308 (1.408) | 0.115 (0.284, 0.686) | -0.018 (0.445, 0.967) | -0.028 (0.414, 0.946) |
| MoCA-J | 25.087 (2.745) | 26.391 (2.231) | 24.654 (3.261) | 25.885 (3.051) | 1.231 (0.526, 0.024) | 0.433 (0.821, 0.600) | 0.074 (0.767, 0.924) |
| Category fluency | 17.478 (3.764) | 18.043 (5.555) | 17.846 (4.730) | 17.115 (4.412) | -0.731 (0.797, 0.364) | -0.368 (1.332, 0.783) | 1.296 (1.164, 0.271) |
| Letter fluency | 14.130 (4.159) | 14.478 (5.846) | 14.808 (3.555) | 14.462 (3.765) | -0.346 (0.642, 0.592) | -0.677 (1.254, 0.591) | 0.694 (0.937, 0.462) |
| Logical memory I (immediate) | 10.000 (4.852) | 11.696 (3.971) | 10.346 (4.039) | 12.346 (4.354) | 2.000 (0.888, 0.029) | -0.346 (1.234, 0.780) | -0.304 (1.296, 0.815) |
| Logical memory II (delayed) | 7.826 (4.783) | 9.696 (4.028) | 9.500 (4.150) | 11.423 (5.085) | 1.923 (0.927, 0.044) | -1.674 (1.299, 0.202) | -0.054 (1.353, 0.969) |
| Symbol | 67.130 (14.169) | 68.000 (15.886) | 68.115 (13.840) | 69.000 (13.291) | 0.885 (1.181, 0.457) | -0.985 (4.088, 0.811) | -0.015 (1.723, 0.993) |
| Digit span forward | 9.478 (1.855) | 10.391 (3.461) | 9.846 (2.738) | 10.269 (2.808) | 0.423 (0.523, 0.423) | -0.368 (0.794, 0.645) | 0.490 (0.763, 0.524) |
| Digit span backward | 7.739 (2.179) | 7.870 (1.984) | 7.577 (1.748) | 8.731 (2.409) | 1.154 (0.364, 0.003) | 0.162 (0.600, 0.788) | -1.023 (0.531, 0.060) |
| TMT-A | 4.402 (0.514) | 4.497 (0.538) | 4.267 (0.504) | 4.213 (0.287) | -0.054 (0.089, 0.550) | 0.135 (0.134, 0.317) | 0.149 (0.130, 0.259) |
| TMT-B | 4.881 (0.595) | 4.856 (0.602) | 4.745 (0.559) | 4.740 (0.523) | -0.004 (0.101, 0.966) | 0.137 (0.163, 0.404) | -0.021 (0.148, 0.886) |
| Brain volumes ($\ell$) | | | | | | | |
| GM | 0.567 (0.048) | 0.561 (0.049) | 0.560 (0.051) | 0.556 (0.048) | -0.004 (0.002, 0.079) | 0.007 (0.014, 0.599) | -0.002 (0.003, 0.412) |
| WM | 0.393 (0.043) | 0.392 (0.045) | 0.391 (0.036) | 0.389 (0.035) | -0.002 (0.001, 0.037) | 0.002 (0.011, 0.834) | 0.001 (0.002, 0.726) |
| CSF | 0.475<br>(0.075) | 0.478<br>(0.073) | 0.453<br>(0.074) | 0.452<br>(0.074) | -0.001<br>(0.002, 0.728) | 0.022<br>(0.021, 0.296) | 0.003<br>(0.003, 0.325) |
| TBV | 0.961<br>(0.077) | 0.953<br>(0.078) | 0.951<br>(0.078) | 0.945<br>(0.074) | -0.006<br>(0.002, 0.001) | 0.010<br>(0.022, 0.658) | -0.002<br>(0.003, 0.474) |
| TIV | 1.436<br>(0.129) | 1.431<br>(0.127) | 1.404<br>(0.120) | 1.397<br>(0.122) | -0.007<br>(0.001, 0.000) | 0.032<br>(0.036, 0.370) | 0.002<br>(0.002, 0.369) |

*Note.* Scores of TMT-A and -B were logarithmic transformed.

**Table S4.** Comparison of levels of biomarkers at pre- and post-experiment between the intervention and control groups.

|  | Intervention |  | Control |  | Estimates |  |  |
| --- | --- | --- | --- | --- | --- | --- | --- |
|  | Pre Mean (SD) | Post Mean (SD) | Pre Mean (SD) | Post Mean (SD) | Time (SE, *p*) | Group (SE, *p*) | Time × Group (SE, *p*) |
| NfL | 13.384 (3.705) | 16.131 (5.473) | 16.312 (6.231) | 18.346 (4.824) | 2.034 (0.729, 0.008) | -2.928 (1.480, 0.052) | 0.714 (1.064, 0.506) |
| Aβ40 | 11.607 (9.379) | 11.059 (6.599) | 10.760 (6.275) | 38.976 (102.496) | 28.215 (14.534, 0.058) | 0.847 (14.194, 0.953) | -28.764 (20.851, 0.174) |
| Aβ42 | 27.544 (29.835) | 20.950 (20.180 ) | 28.323 (30.963) | 17.917 (25.217) | -10.781 (6.533, 0.106) | -0.779 (7.745, 0.920) | 3.986 (9.477, 0.676) |
| log(NfL) | 2.556 (0.284) | 2.734 (0.306) | 2.732 (0.341) | 2.876 (0.268) | 0.143 (0.035, 0.000) | -0.176 (0.086, 0.046) | 0.035 (0.051, 0.500) |
| log(Aβ40) | 2.199 (0.719) | 2.208 (0.689) | 2.221 (0.564) | 2.143 (1.485) | -0.084 (0.247, 0.735) | -0.023 (0.262, 0.931) | 0.088 (0.354, 0.806) |
| log(Aβ42) | 2.949 (0.780) | 2.702 (0.837) | 2.908 (0.896) | 2.271 (1.156) | -0.651 (0.202, 0.002) | 0.042 (0.266, 0.876) | 0.395 (0.293, 0.185) |

*Note.* For Aβ40, two participants and five ones were excluded from the intervention and control groups at the endpoint due to lower levels than the lower limit of the measurable range. For Aβ42, one participant was excluded from the intervention group at the endpoint due to the level lower than the measurable range, and one participant and another participant in the control group were excluded due to levels lower and higher than the measurable range.

